# The Influence Of Covid-19 On Patient Mobilization And Injury Attributes In The ICU: A Retrospective Analysis Of A Level II Trauma Center

**DOI:** 10.1101/2023.10.25.23297544

**Authors:** Yelissa Navarro, Elizabeth Huang, Chandler Johnson, Forrest Clark, Samuel Coppola, Suraj Modi, Gordon L. Warren, Jarrod A. Call

## Abstract

The objectives of this study were to determine the effect of COVID-19 on physical therapy (PT) mobilization of traumatically-injured patients and to determine if mobilization affected patient course in the ICU. This retrospective study included patients who were admitted to the ICU of a level II trauma center. The patients were divided into two groups, i.e., those admitted before (n=378) and after (n=499) April 1, 2020 when Georgia’s COVID-19 Shelter-in-place order was mandated. The two groups were contrasted on nominal and ratio variables using Chi-square and Student’s t-tests. A secondary analysis focused specifically on the after COVID patients examined the extent to which mobilization (n=328) or lack of mobilization (n=171) influenced ICU outcomes (e.g., mortality, readmission). The two groups were contrasted on nominal and ratio variables using Chi-square and Student’s -tests. The after COVID patients had higher injury severity as a greater proportion was classified as severely injured (i.e., >15 on Injury Severity Score) compared to the before COVID patients. After COVID patients also had greater cumulative number of comorbidities and experienced greater complications in the ICU. Despite this, there was no difference between patients in receiving a PT consultation or day-to-mobilization. Within the after COVID cohort, those that were mobilized were older, a higher proportion were female, they had greater Glasgow Coma Scale scores, had longer total hospital days, and a lesser mortality rate. Despite shifting patient injury attributes post-COVID-19, a communicable disease, mobilization care remained consistent and effective.

**Level of Evidence:** Level III

## INTRODUCTION

In 2020, due to the coronavirus pandemic caused by SARS-CoV-2 (COVID-19), the United States was forced to enact sudden measures to help alleviate the spread of the virus. A ‘shelter-in-place’ approach was executed, encouraging persons to stay at home, and many large gatherings were canceled. Travel was strictly controlled, especially international, and a variety of protective public health initiatives were put forward, namely an emphasis on handwashing, mask wearing, and social distancing with ‘stay-at-home’ orders [1]. Hospitals experienced a nationwide drop in overall admissions beginning in March of 2020, including acutely ill non-COVID-19 patients [2]. It has been theorized that a large reason for this was the avoidance of seeking hospital care by patients, mainly in response to media messaging and hesitancy to leave home with ‘shelter-in-place’ orders in effect [2, 3]. With these restrictions in mind, we sought to assess the pandemic’s effect on intensive care unit (ICU) trauma admissions and care.

In the treatment of critically ill patients, early mobilization and early physical therapy (PT) intervention have been associated with a reduced stay in the ICU [4–13], reduced ICU healthcare costs [5, 6, 11, 14–17], reduced length of stay in the hospital [5, 12–14, 18–25], and decreased hospital readmissions [4]. Additionally, improved patient functional outcomes and decreased need for care after hospital discharge have been associated with early mobilization (2, 4-7, 9-12, 16-23). However, PT and mobilization require close human interaction, and after March 2020, these interactions were taking place within the global context of a highly-communicable disease. Therefore, the first objective of our study was to determine whether the onset of COVID-19 was associated with a change in the percentage of patients receiving a PT mobilization order and time to mobilization. The second objective of our study was to determine if there were differences in patient characteristics, injury attributes, hospital stay, and readmission rates for patients in the “after COVID onset” group that did or did not receive mobilization in the ICU.

## METHODS

This was a retrospective analysis of traumatically-injured patients that were admitted to the ICU at Piedmont Athens Regional Medical Center (PARMC) in Athens, Georgia. PARMC is a 360-bed, non-profit hospital that offers a level II trauma center and serves Athens and 17 counties in Georgia. Approval for this study was granted by the Piedmont Institutional Review Board and Piedmont Athens Regional, IRB #: 1751170-1 Piedmont.

This study specifically looked at patients admitted to the ICU from the emergency room at PARMC due to traumatic injury. Our analysis included patients who were admitted from January 1, 2019, to December 31, 2021, and patients were selected using inclusion criteria outlined by the National Trauma Data Standard (NTDS). Emergency room patients were first entered in the trauma registry if they fulfilled NTDS inclusion criteria. Our study then selected for patients who were recorded in the trauma registry and were subsequently admitted to the ICU. Patients meeting these criteria were identified in the hospital’s trauma registry, a database that provided detailed information including demographics, mechanisms of injury, and patient outcomes. In total, 877 patients met the criteria for our analysis.

The time period for data collection in our study encompassed the COVID-19 pandemic and the shutdowns that followed in its wake. Patients were divided into two groups: “before COVID onset” (n= 378) and “after COVID onset” (n= 499), meaning after the onset of COVID-19. To demarcate these two groups, the threshold date for the COVID-19 pandemic was defined as April 1, 2020, with patients admitted to the ICU on or before March 31, 2020, classified as “before COVID onset” and those admitted on or after April 1, 2020, as “after COVID onset”. Any patients that were discharged less than 24 hours after admission, downgraded to another unit less than 24 hours after ICU admission, transferred to another institution, or who expired less than 24 hours after admission were not included in the patient group. Analyses were then performed to look for differences in patient and injury characteristics as well as changes in care, such as PT consultation, in the “before COVID onset” and “after COVID onset” settings. A secondary analysis was eventually conducted to determine patient characteristics and outcomes within the “after COVID onset” group that were mobilized (Mobilized) or were not mobilized (Not Mobilized).

To analyze differences in patient outcomes, clinical data were extracted from the hospital’s trauma registry and patient charts. The variables extracted consisted of patient demographics, injury types, chief complaints, injury severity scores, hospital course, PT consultation, and time to mobilization. Patient demographics were defined as: age, sex (male or female), ethnicity (Hispanic/Latino or non-Hispanic/Latino), and race [White/Caucasian or Other (Black/African-American, Asian, Hispanic, American-Indian/Alaskan Native, Pacific Islander, or refused to answer)]. Injury types were defined as blunt or penetrating according to the descriptions of trauma types in AOTR Alliance of Trauma Registry Resource Manual [26]. Chief complaints were categorized as: falls, gunshot wounds, motor vehicle crash, or other (assault, bicycle, burn, knife, pedestrian, puncture wound, other blunt mechanism, other penetrating mechanism, and unknown). Recorded vitals upon ER admission were weight, height, and body mass index. Injury severity scores included: Glasgow Coma Score (GCS), Injury Severity Score (ISS), severe ISS (scores greater than 15), New Injury Severity Score (NISS), Trauma and injury severity scores (TRISS), and the Revised Trauma Score (RTS) (see **Supplemental Appendix Table 1**). Hospital course of care data included: day of the week patient was admitted to the ER, length of stay in the ICU, length of stay in the hospital itself, days spent on the ventilator, and readmission. PT was indicated on the data record sheet if the patient received a PT consultation and was later mobilized. Time to mobilization was recorded as the number of days it took for the patient to be mobilized by PT from the day of admission. For readmissions, data that were recorded were the number of days post-ED admission before the patient was readmitted and the number of days patients were readmitted.

**Table 1.** Patient characteristics stratified by before and after COVID onset.

|  |  |  |  |
| --- | --- | --- | --- |
| Patient Characteristics | Before COVID onset<br>(n=378) | After COVID onset<br>(n=499) | P value |
| Age at Admission, years (IQR) | 50 (42.25) | 55 (40) | 0.70 |
| Gender, n (%) |  |  | 0.86 |
| Male | 244 (64.6%) | 325 (65.1%) |  |
| Female | 134 (35.4%) | 174 (34.9%) |  |
| Ethnicity, n (%) |  |  | 0.46 |
| Hispanic or Latino | 16 (4.0%) | 24 (4.8%) |  |
| Not Hispanic or Latino | 361 (96.0%) | 475 (95.2%) |  |
| Race, n(%) |  |  | 0.016 |
| White or Caucasian | 305 (80.7%) | 368 (73.8%) |  |
| Other (Asian, Black/African American, Native Hawaiian, Pacific Islander, Refused) | 73 (19.3%) | 131 (26.2%) |  |
| Vitals |  |  |  |
| Height, meters (IQR) | 1.73 (0.15) | 1.73 (0.15) | 0.50 |
| Weight, kilograms (IQR) | 79.9 (27.03) | 80.45 (29.88) | 0.91 |
| Body Mass Index (IQR) | 26.89 (8.58) | 26.65 (8.40) | 0.74 |
| Systolic Blood Pressure (IQR) | 132 (36.25) | 132 (34) | 0.30 |
| Diastolic Blood Pressure (IQR) | 81 (22.25) | 81 (22) | 0.43 |
| Pulse (IQR) | 89 (29) | 87 (30) | 0.69 |
| Respiratory Rate (IQR) | 18 (4) | 18 (6) | 0.91 |
| SpO<sub>2</sub> (IQR) | 97 (5) | 97 (4) | 0.27 |
| Injury Type, n (%) |  |  | 0.29 |
| Blunt | 349 (92.3%) | 447 (89.6%) |  |
| Penetrating | 29 (7.7%) | 51 (10.2%) |  |
| Other (Thermal, not applicable) | 0 | 1 (0.2%) |  |
| Chief Complaint, n (%) |  |  | 0.046 |
| Fall | 161 (42.6%) | 176 (35.3%) |  |
| Gunshot wound | 17 (4.5%) | 41 (8.2%) |  |
| Motor vehicle crash | 177 (46.8%) | 250 (50.1%) |  |
| Other (Assault, Bicycle, Burn, Knife, Pedestrian, Puncture wound, Other blunt mechanism, Other penetrating mechanism, Struck by object or person, Unknown) | 23 (6.1%) | 32 (6.4%) |  |
| Injury Severity |  |  |  |
| Severe ISS, n (%) | 177 (46.83%) | 292 (58.52%) | 0.001 |
| TRISS (IQR) | 0.968 (0.044) | 0.965 (0.068) | 0.014 |
| ISS (IQR) | 14 (13) | 17 (15) | < 0.001 |
| NISS (IQR) | 17 (16) | 22 (16) | < 0.001 |
| RTS (IQR) | 12 (1) | 12 (1) | 0.21 |
| *IQR=interquartile range |  |  |  |

Other important variables that were extracted included comorbidities, complications, and mortality. Comorbidities recorded from charts were alcohol-use disorder, drug-use disorder, currently receiving chemotherapy, congenital anomalies, congestive heart failure, smoker, chronic renal failure, previous cerebrovascular accident, diabetes, currently has cancer, possesses an advanced directive, inability to perform activities-of-daily-living, previous history of angina, previous history of myocardial infarction, peripheral vascular disease, hypertension, born prematurely, chronic obstructive pulmonary disease, steroid-use, liver cirrhosis, dementia, attention-deficit/hyperactivity disorder, and any other major psychiatric illness not mentioned, any other disorders not mentioned. Complications recorded were deep surgical infection, previous history of drug/ alcohol use disorder development, deep venous thrombosis, compartment syndrome, graft/prosthesis/flap failure, myocardial infarction, organ space infection, osteomyelitis, pneumonia, pulmonary embolism, sepsis, stroke/cerebrovascular accident, superficial infection, unplanned intubation, unplanned return to operating room, and urinary tract infection. Mortality was recorded as whether the patient was discharged alive or expired.

Data were extracted in two stages with the goal of determining whether patient mobilization following a traumatic injury was affected by the onset of COVID. In the first stage, all trauma patients that that were admitted to the emergency room and fulfilled the NTDS criteria were recorded on the PARMC trauma registry. This trauma registry included mechanisms of injury, injury code, hospital stay duration, where the patient was discharged afterwards, and the ICD-10 diagnosis of, at maximum, six of the patients’ injuries. In the second stage, patient medical records were analyzed for all patients on the trauma registry that were then admitted to the ICU. During this chart review, each patient record was analyzed by two independent investigators to extract and confirm the variables listed above, and to confirm the previously extracted data from the trauma registry.

All patients were de-identified during the statistical analysis phase. The two groups (e.g., “before COVID onset” and “after COVID onset”) were compared using Mann-Whitney tests because the datasets were not normally distributed. Results are reported as median and interquartile range. The two groups were contrasted on nominal variables using Chi-square tests. All statistical analyses were performed using JMP statistical software (JMP 16, SAS, Cary, NC USA).

## RESULTS

### “Before and after COVID onset” patient characteristics, injury attributes, and mobilization

For the 877 patients, demographic, basic clinical measures, and patient injury types that were ratio data are summarized in **Table 1**. Median patient age was not different between the two groups (i.e., the “before COVID onset” and “after COVID onset” groups) (50 vs. 55 years, p=0.70). There was a significant race distribution difference such that Other (Asian, Black/African American, Native Hawaiian, Pacific Islander) represented a greater proportion of patients in the “after COVID onset” group vs. the “before COVID onset” group (26% vs. 19%, p=0.016). There were no statistically significant differences in basic clinical measures (e.g., height, weight, body mass index) between groups (**Table 1**).

Blunt trauma was the most common injury type, independent of group, and this was reflected within the chief complaint data as high proportions for complaints identified as falls (38% of total) and motor vehicle accidents (49% of total). There was a statistically significant chief compliant distribution shift such that gunshot wounds represented a greater proportion of complaints in the “after COVID onset” group vs. the “before COVID onset” group (8.2% vs. 4.5%, p=0.046). Overall, a higher proportion of “after COVID onset” patients were classified as severely injured, i.e., an ISS greater than 15, compared to “before COVID onset” patients (58.5% vs. 46.8%, p<0.001). In addition, ISS and NISS scores were, on average, significantly higher in the “after COVID onset” patients (**Figure 1A**). TRISS scores, which estimate the survivability of patients, were significantly less in the “after COVID onset” patients (**Figure 1A**). Finally, “after COVID onset” patients presented to the ICU with greater total comorbidities and experienced greater total complications while in the ICU (**Figure 1B-C**).

**Figure 1:**
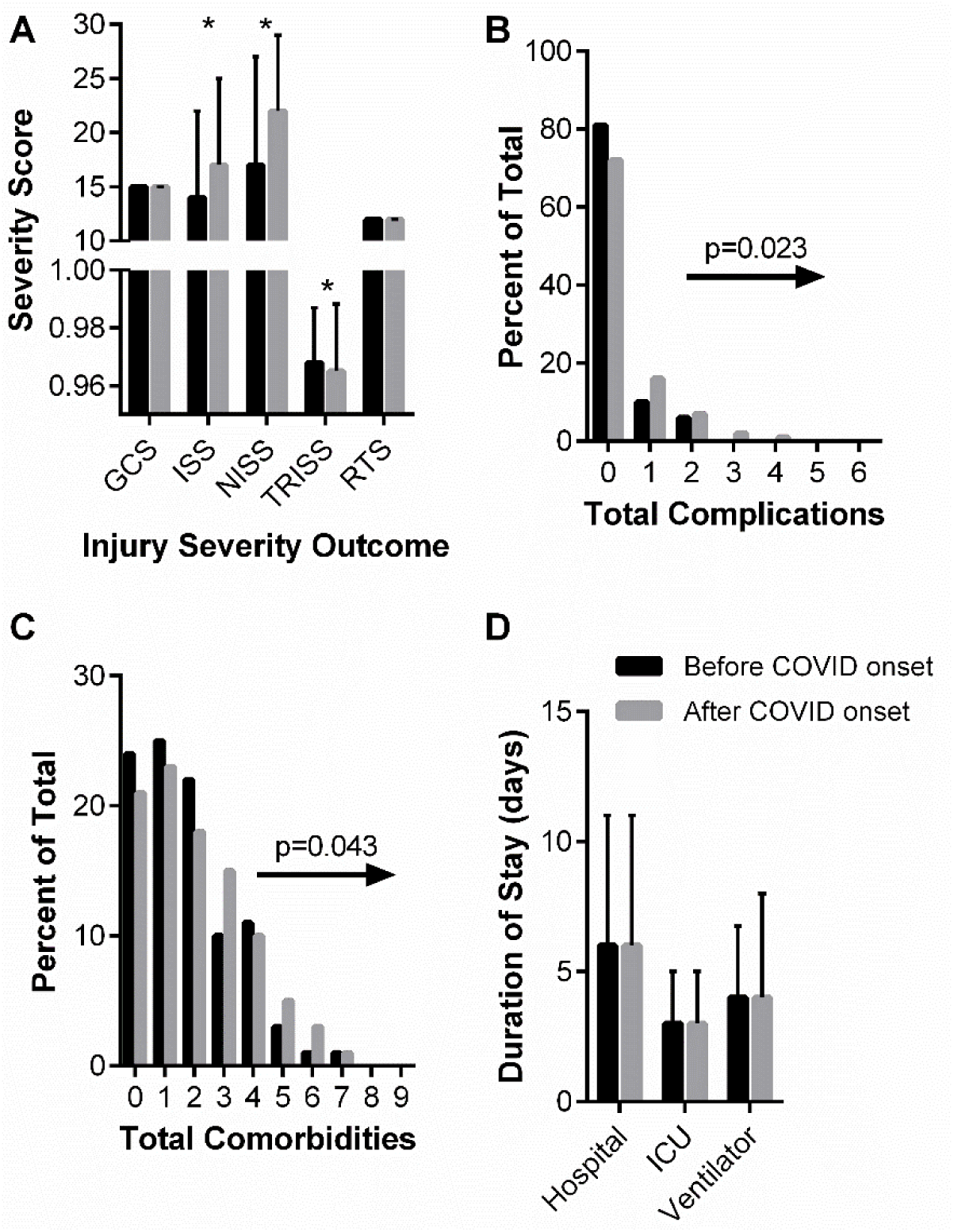
“before and after COVID onset” patient injury severity (A), cumulative complications (B), cumulative comorbidities (C), and hospital durations (D). (A) Glasgow Coma Score (GCS), Injury Severity Score (ISS), New Injury Severity Score (NISS), Trauma Score and Injury Severity Score (TRISS), and Revised Trauma Score (RTS) are shown as median and interquartile range. Data analyzed using Mann Whitney with * representing statistical difference between groups (p<0.05). (B) Chi-squared analysis of distribution shifts of total patient complications between “before COVID onset” and “after COVID onset” cohorts. Statistical p-value and distribution shift direction is indicated. (C) Chi-squared analysis of distribution shifts of total patient comorbidities between “before COVID onset” and “after COVID onset” cohorts. Statistical p-value and distribution shift direction is indicated. (D) Hospital, ICU, and ventilator durations are shown as median and interquartile range. Data analyzed using Mann Whitney.

Individual comorbidities and complications for the before- and “after COVID onset” analysis are reported in **Supplemental Tables 1** and **2**, respectively. A greater percentage of “after COVID onset” patients presented to the ICU with disseminated cancer (6% vs. <1%, p<0.001) and advance directives limiting care (∼13% vs. 2%, p<0.001) compared to the “before COVID onset” patients. There were statistical trends for prior cerebral vascular accidents, steroid use, and other major psychiatric disorders. For complications arising during the hospital course, only stroke/cerebral vascular accident was statistically different between patient groups.

**Table 2** summarizes patient hospital courses stratified by “before COVID onset” and “after COVID onset” patients. The day of the week patients were admitted (weekends vs. weekdays) remained similar between the two groups. Important to the primary objective for this retrospective analysis, there were no statistical differences in the percentage of patients receiving PT nor the median day to mobilization in the ICU between before- and “after COVID onset” groups (**Table 2**). Approximately 65% of patients received a PT order and the median time to mobilization was 4 days “before COVID onset” and 3 days “after COVID onset” (p=0.86). The lengths of stay in the hospital, in the ICU, and on a ventilator did not significantly differ before- vs. “after COVID onset” (**Figure 1D**). In agreement with “after COVID onset” patients having greater injury severity, a lower predicted survivability, and greater total complications, there was also a greater mortality associated with the “after COVID onset” patients, going from 4.8% mortality in the “before COVID onset” group to 10.7% mortality in the “after COVID onset” group (p=0.001). “after COVID onset” patients were also more likely to be readmitted (14% vs. 6%, p<0.001) compared to “before COVID onset” patients.

**Table 2.** Hospital stay characteristics stratified by before and after COVID onset.

|  |  |  |  |
| --- | --- | --- | --- |
| Hospital Stay Characteristics | Before COVID onset<br>(n= 378) | After COVID onset<br>(n= 499) | P value |
| Weekend vs. Weekday Admission, n (%) |  |  | 0.66 |
| Weekday | 263 (69.6%) | 354 (70.9%) |  |
| Weekend | 115 (30.4%) | 145 (29.1%) |  |
| Day of the Week Admission, n (%) |  |  | 0.51 |
| Sunday | 48 (12.7%) | 71 (14.2%) |  |
| Monday | 57 (15.1%) | 71 (14.2%) |  |
| Tuesday | 53 (14.0%) | 58 (11.6%) |  |
| Wednesday | 42 (11.1%) | 75 (15.0%) |  |
| Thursday | 56 (14.8%) | 72 (14.4%) |  |
| Friday | 55 (14.6%) | 78 (15.6%) |  |
| Saturday | 67 (17.7%) | 74 (14.8%) |  |
| Received PT, n (%) | 215 (56.9%) | 302 (60.6%) | 0.97 |
| PT Time to Mobilization, day (IQR) | 4 (4) | 3 (3.25) | 0.86 |
| Discharge Status, n (%) |  |  | 0.001 |
| Alive | 360 (95.2%) | 445 (89.2%) |  |
| Expired | 18 (4.8%) | 54 (10.8%) |  |
| Readmission |  |  |  |
| Patients Readmitted, n (%) | 23 (6.08%) | 70 (14.03%) | 0.0003 |
| Days Post ED Before Readmission, days (IQR) | 23.92 (37.47) | 42.00 (83.21) | 0.04 |
| Length of Readmission Stay, days (IQR) | 5 (10) | 2 (3.92) | 0.03 |
| *IQR=interquartile range |  |  |  |

### “after COVID onset” patient characteristics, injury attributes and outcomes stratified by mobilization

Our secondary analysis examined patient attributes and hospital courses within the “after COVID onset” group based on whether or not patients were mobilized. Of the 499 “after COVID onset” patients, those that were mobilized were older (median age: 57 vs. 50 years, p=0.005), female (74% female Mobilized vs. 68% male Mobilized, p=0.004), present with a Blunt Injury, and not have sustained a gunshot wound (**Table 3**). In fact, patients that sustained a gunshot wound were 55% less likely to be mobilized compared to patients classified with chief complaints of falls and motor vehicle crashes. There were no statistical differences between patient groups for basic clinical outcomes (**Table 3**).

**Table 3.** Patient characteristics after-COVID stratified by mobilization.

|  |  |  |  |
| --- | --- | --- | --- |
| Patient Characteristics | Not Mobilized<br>(n=171) | Mobilized<br>(n=328) | P value |
| Age at Admission, years (IQR) | 50 (38) | 57 (41) | 0.005 |
| Gender, n (%) |  |  | 0.004 |
| Male | 126 (73.7%) | 199 (60.7%) |  |
| Female | 45 (26.3%) | 129 (39.3%) |  |
| Ethnicity, n (%) |  |  | 0.57 |
| Hispanic or Latino | 6 (3.5%) | 13 (4.0%) |  |
| Not Hispanic or Latino | 164 (96.5%) | 309 (96.0%) |  |
| Race, n(%) |  |  | 0.51 |
| White or Caucasian | 123 (71.9%) | 245 (74.7%) |  |
| Other (Asian, Black/African American,<br>Native Hawaiian, Pacific Islander, Refused) | 48 (28.1%) | 83 (25.3%) |  |
| Vitals |  |  |  |
| Height, meters (IQR) | 1.75 (0.15) | 1.727 (0.17) | 0.071 |
| Weight, kilograms (IQR) | 81.6 (29.7) | 79.5 (30.7) | 0.95 |
| Body Mass Index (IQR) | 26.75 (8.81) | 26.63 (8.32) | 0.57 |
| Systolic Blood Pressure (IQR) | 131 (47) | 132.5 (32.49) | 0.78 |
| Diastolic Blood Pressure (IQR) | 81 (24) | 81 (21.75) | 0.091 |
| Pulse (IQR) | 88 (27) | 86.5 (32) | 0.25 |
| Respiratory Rate (IQR) | 18 (6) | 18 (6) | 0.79 |
| SpO<sub>2</sub> (IQR) | 98 (5) | 97 (4) | 0.33 |
| Injury Type, n (%) |  |  | <0.001 |
| Blunt | 141 (82.4%) | 305 (93.3%) |  |
| Penetrating | 30 (17.5%) | 21 (6.4%) |  |
| Other (Thermal, not applicable) | 0 (0%) | 1 (0.3%) |  |
| Chief Complaint, n (%) |  |  | <0.001 |
| Fall | 53 (30.1%) | 123 (37.5%) |  |
| Gunshot wound | 26 (15.2%) | 15 (4.6%) |  |
| Motor vehicle crash | 76 (44.4%) | 174 (53.0%) |  |
| Other (Assault, Bicycle, Burn, Knife,<br>Pedestrian, Puncture<br>wound, Other blunt mechanism, Other<br>penetrating mechanism, Struck by object<br>or person, Unknown) | 16 (9.4%) | 16 (4.9%) |  |
| Injury Severity |  |  |  |
| Severe ISS, n (%) | 99 (33.9%) | 193 (66.1%) | <0.001 |
| TRISS (IQR) | 0.97 (0.0985) | 0.97 (0.056) | 0.27 |
| ISS (IQR) | 17 (16) | 17 (14) | 0.73 |
| NISS (IQR) | 22 (22) | 22 (15) | 0.028 |
| RTS (IQR) | 12 (2) | 2 (0.75) | <0.001 |
| *IQR=interquartile range |  |  |  |

In regards to patient injury severity, the proportion of mobilized patients classified as severely injured was not statistically different from the proportion of patients not mobilized (∼58%); however, mobilized patients had greater GCS and RTS scores, and lower NISS scores (**Figure 2A**). GCS is a predictive coma score such that a ‘15’ is normal and lower scales are associated with a greater likelihood of coma, vegetative state, and ultimately death. The RTS is a trauma score that correlates statistically with greater survival. There were no significant differences between patient groups for total comorbidities presented in the ICU nor total complications experience within the ICU (**Figure 2B-C**).

**Figure 2:**
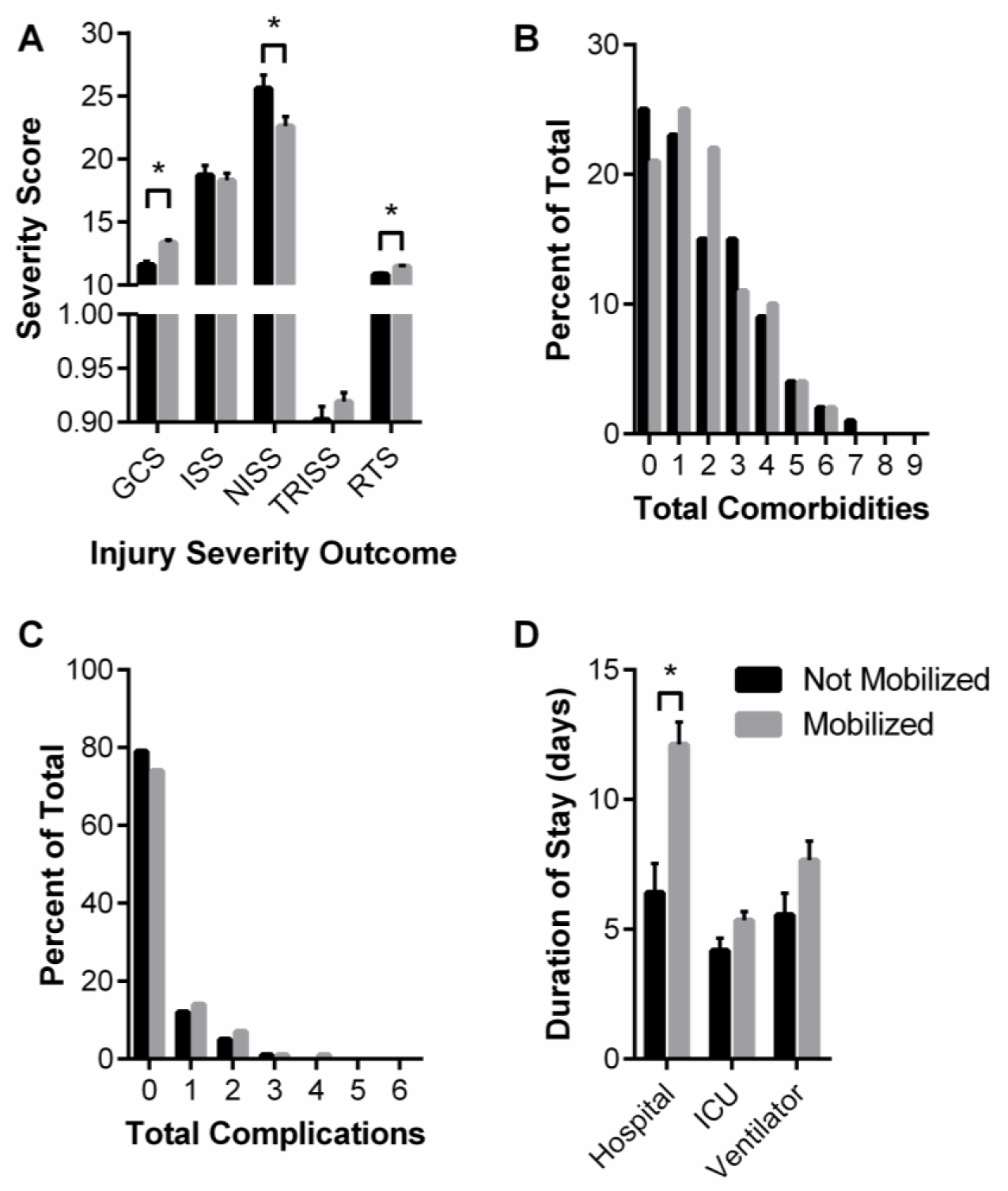
“after COVID onset” mobilized and not mobilized patient injury severity (A), cumulative complications (B), cumulative comorbidities (C), and hospital durations (D). (A) Glasgow Coma Score (GCS), Injury Severity Score (ISS), New Injury Severity Score (NISS), Trauma Score and Injury Severity Score (TRISS), and Revised Trauma Score (RTS) are shown as median and interquartile range. Data analyzed using Mann Whitney with * representing statistical difference between groups (p<0.05). (B) Chi-squared analysis of distribution shifts of total patient complications. (C) Chi-squared analysis of distribution shifts of total patient comorbidities. (D) Hospital, ICU, and ventilator durations are shown as median and interquartile range. Data analyzed using Mann Whitney with * representing statistical difference between groups (p<0.05.

Individual comorbidities and complications for the Mobilized vs. Not Mobilized groups are reported in **Supplemental Tables 3** and **4**, respectively. A lesser percentage of Mobilized patients presented to the ICU with advance directive limiting care (∼6% vs. 24%, p<0.001) compared to the Not Mobilized patients. There were statistical trends for hypertension and cirrhosis. For complications arising during the hospital course of care, there were only statistical trends for myocardial infarctions and pulmonary embolism.

**Table 4** summarizes patient hospital courses stratified by “after COVID onset” patient mobilization. The day of the week and weekday vs. weekend was not associated with whether or not a patient was mobilized. Interestingly, with respect to patient outcomes, patients who were mobilized were more likely to be discharged alive (98% vs. 72%, p<0.001), although they are also more likely to be readmitted (18% vs. 6%, p<0.001). Mobilized patient’s median hospital stay was greater (8 vs. 3 days, p<0.001), but ICU duration and days on a ventilator were not different between the groups (**Figure 2D**).

**Table 4.**
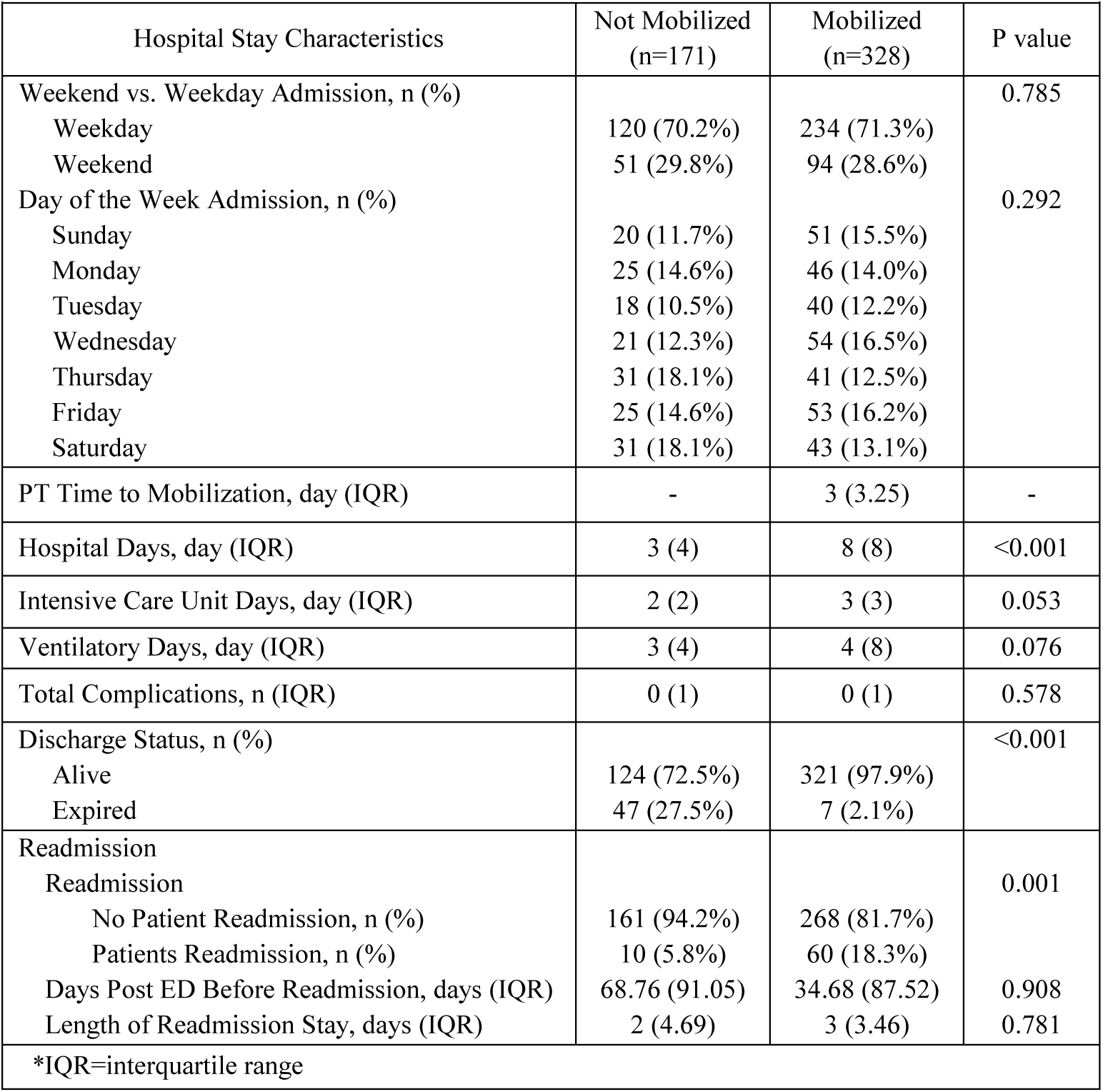
Hospital stay characteristics after-COVID stratified by PT and no PT.

## DISCUSSION

Our retrospective study revealed that there was an association between the onset of COVID-19 and ICU patients with greater injury severity, complications, comorbidities, and mortality rate. There was also evidence that the types of injury patients presented to the ICU shifted after the onset of COVID-19, in agreement with other studies [27, 28]. Despite these changes, we report herein that there was no difference in the proportion of patients receiving a PT consult or time to mobilization for patient groups “before COVID onset” vs “after COVID onset”. Patient injury characteristics notwithstanding, this is a significant finding considering the early uncertainty of the communicable COVID-19 disease.

In the two years following the start of the COVID-19 pandemic, the most significant change seen among the different groups was a rise in the severity of trauma injury requiring patient admission to the ICU. The consensus among the literature suggests the main cause of these findings to be a delay in patients seeking care due to fear of COVID-19 infection [29, 30]. Herein, the proportion of patients classified as “severely injured” was greater in the “after COVID onset” group, and additionally, we detected total comorbidities shifts suggesting patients were delaying their care to avoid any healthcare setting. Of the twenty-three comorbidities we tracked, it is notable that patients presenting with disseminated cancer was greater in the “after COVID onset” group (**Supplemental Table 1**). We ran ANCOVAs to control for either cumulative patient complications or disseminated cancer, and there was still no significant difference between “before COVID onset” and “after COVID onset” groups in days to mobilization (p≥0.779). This retrospective analysis suggests that the PT staff at this level II trauma center were able to manage the shift in patient injury attributes and complications. The extent to which these results would be similar to a level I trauma center are unclear.

Recent studies have attempted to capture changes in patient characteristics of traumatically-injured ICU admissions after the COVID-19 pandemic forced adjustment to the lives of populations around the globe [31–34]. In a large, multicenter retrospective study that analyzed the first three months of the stay-at-home order, Yeates et al. found that “after COVID onset” trauma patients had decreased lengths of stay in both ICU and total hospital days [31]. These findings are mirrored among other studies that looked at the changes in admission characteristics in the time period immediately following the installment of the stay-at-home orders [35–37]. The results of this study show that there were no major changes in patient demographics; however, the types of injuries resulting in ICU admittance did shift. There was a greater percentage of gunshot wounds as a chief complaint, which is similar to findings reported by others [27, 28].

This study also sought to investigate the relationship between the COVID-19 pandemic and physical therapy consultation. Our analysis of “after COVID onset” patients exclusively that did or did not receive mobilization revealed that mobilized patients had a greater hospital duration compared to those that were not mobilized. Previous experimental studies found early mobility in the ICU to be associated with fewer days on mechanical ventilation [38] and fewer days spent both in the ICU and in the hospital [5–11, 13, 39–41]. It is possible that the effectiveness of the mobilization protocol differed between this retrospective study and the experimental studies where the protocol’s implementation could be tightly controlled. It is also possible that the retrospective vs. experimental studies difference in outcomes could be due to differing patient characteristics such as injury severity, case management decisions such as discharge disposition, differences in physician decisions regarding extubation, or perhaps the level of mobilization attained by the patients.

In the last few decades, viral pandemics that have had global effects have been met with major strategies to combat the spread among the population. The COVID-19 pandemic has been the most recent and has had the most significant response on a global scale involving major social, healthcare, and international changes [46]. Whether or not these changes to the characteristics of patient populations are permeant remain to be elucidated. Research on the H1N1 influenza pandemic of 2009, which was the most recent prior to Sars-CoV-2 pandemic, describes various impacts on healthcare institutions including overwhelming EDs and changes to staffing to overcome pandemic surges that have highlighted the need for institutional preparedness [47]. Health institutional changes from the lessons learned during the previous pandemics and during the time period immediately following the start of the COVID-19 pandemic including changes to length-of-stay, time to transfer and time to intervention for non-COVID traumatically injured patients, early medical interventions including antivirals, and supportive therapies to alleviate milder symptoms As such, hospitals and healthcare institutions may require a shift in expectations of potential trauma admissions up to two years after a global pandemic. This may involve the formation of guidelines in addition to pandemic preparedness for the reallocation of resources to adjust for switch in more severely injured patients during and in the years after a pandemic including availability of PT, further training of personnel, and hospital resources to meet the need of more severely injured trauma patients.

Limitations of this study include its retrospective nature that relies on accurate documentation and could be influenced by charting discrepancies. Additionally, retrospective analysis cannot determine causation of the reported findings and can only evaluate associations or relationships among the patient characteristics and outcomes. The focus on cases from a single institution may lack external validity. As such, future studies should focus on the long-term effects of COVID-19 on regional and national levels. Furthermore, the limitations of the hospital in this study, including its Level-II status, potentially involve the transfer of more severe injuries to outside hospitals which may skew the reproducibility among hospitals of different capacities and patient populations.

## Data Availability

All data produced in the present study are available upon reasonable request to the authors

## ACKNOWLEDGEMENTS

Data for this project was obtained using the Trauma Registry Database at PARMC. Special thanks to Heather Morgan and the Trauma Department at PARMC who provided access to the registry for their hospital. The content is solely the responsibility of the authors and does not necessarily represent the official views of PARMC.

