## Supplemental Appendix 1 for "The Influence Of Covid-19 On Patient Mobilization And Injury Attributes In The ICU: A Retrospective Analysis Of A Level II Trauma Center"

**Appendix Tables**

| **Appendix Table 1. Injury Severity Score Definition and Predictive Utility^8^** | | |
| --- | --- | --- |
| **Severity Scoring System** | **Determining Variables** | **Utility** |
| Glasgow Coma Scale (GCS)  Scale 3 (High Severity) to 15 (Low severity) | Eye response, verbal response, motor response | High score correlate with better neurological function |
| Revised Trauma Score (RTS)  Scale 0 (High Severity) to 12 (Low severity) | GCS, Systolic pressure, Respiratory rate | High score correlate with greater survival rate and better outcome |
| Injury Severity Score (ISS)  Scale 1 (low severity) to 75 (high severity) | Top 3 Body Region Injuries | High scores correlate with low probability of survival |
| New Injury Severity Score (NISS)  Scale 1 (low severity) to 75 (high severity) | Top 3 Injuries Regardless of Body Region | High scores correlate with low probability of survival |
| Trauma and Injury Severity Score (TRISS)  Scale 0 (high severity) to 1 (low severity) | RTS and ISS | High scores correlate with high probability of survival |
