## Supplemental Tables 1-4 for "The Influence Of Covid-19 On Patient Mobilization And Injury Attributes In The ICU: A Retrospective Analysis Of A Level II Trauma Center"

| **Supplemental Table 1. Comorbidities by before and after COVID onset** | | | |
| --- | --- | --- | --- |
| Comorbidities (%) | Before COVID onset  (n= 378) | After COVID onset  (n= 499) | P value |
| Total comorbidities (IQR) | 1 (2.25) | 2 (2) | 0.0067 |
| Currently receiving chemotherapy  Congenital anomalies  Congestive Heart Failure  Smoker  Renal failure  Prior cerebral vascular accident  Diabetes  Disseminated cancer  Advanced directive limiting care  History of angina  Prior myocardial infarction  Peripheral vascular disease  Hypertension  Prematurity  COPD  Steroid use  Cirrhosis  Other major non-psychiatric disorder  EtOH use disorder  Other substance use disorder  Dementia  ADHD  Other major psychiatric disorder | 2 (0.53%)  3 (0.79%)  29 (7.67%)  91 (24.07%)  29 (7.67%)  23 (6.08%)  77 (20.37%)  2 (0.53%)  8 (2.12%)  1 (0.26%)  16 (4.23%)  19 (5.03%)  161 (42.59%)  1 (0.26%)  30 (7.94%)  0 (0%)  9 (2.38%)  1 (0.27%)  37 (9.79%)  41 (10.85%)  19 (5.03%)  10 (2.65%)  65 (17.20%) | 5 (1.00%)  2 (0.40%)  52 (10.42%)  136 (27.25%)  45 (9.02%)  48 (9.62%)  83 (16.63%)  30 (6.01%)  63 (12.63%)  1 (0.20%)  31 (6.21%)  28 (5.61%)  204 (40.88%)  0 (0%)  35 (7.01%)  5 (1.00%)  12 (2.40%)  4 (0.8%)  59 (11.82%)  68 (13.63%)  18 (3.61%)  9 (1.80%)  112 (22.44%) | 0.44  0.44  0.16  0.29  0.48  0.057  0.16  <0.0001  <0.0001  0.84  0.20  0.70  0.61  0.25  0.61  0.051  0.98  0.30  0.34  0.22  0.30  0.40  0.0551 |

Supplemental Tables 1 -4

| **Supplemental Table 3. Complications before and after COVID onset** | | | |
| --- | --- | --- | --- |
| Complications (%) | Before COVID onset  (n= 378) | After COVID onset  (n= 499) | P value |
| Total complications (IQR) | 0 (0) | 0 (1) | 0.0046 |
| Deep surgical infection  Drug/EtOH substance use disorder  Deep venous thrombosis (DVT)  Compartment syndrome  Graft/ prosthesis/ flap failure  Myocardial infarction  Organ space infection  Osteomyelitis  Pneumonia  Pulmonary embolism  Sepsis  Stroke/cerebral vascular accident  Superficial infection  Urinary tract infection | 4 (1.06%)  4 (1.06%)  15 (3.97%)  1 (0.26%)  0 (0%)  0 (0%)  0 (0%)  4 (1.06%)  43 (11.38%)  3 (0.79%)  16 (4.23%)  2 (0.53%)  3 (0.79%)  11 (2.91%) | 5 (1.00%)  8 (1.06%)  21 (4.21%)  2 (0.40%)  1 (0.20%)  2 (0.40%)  2 (0.40%)  1 (0.20%)  58 (11.62%)  7 (1.40%)  23 (6.81%)  12 (2.40%)  5 (1.00%)  14 (2.81%) | 0.93  0.49  0.86  0.73  0.38  0.22  0.22  0.095  0.91  0.40  0.10  0.028  0.75  0.93 |

| **Supplemental Table 3. Comorbidities after-COVID stratified by mobilization** | | | |
| --- | --- | --- | --- |
| Comorbidities (%) | Not Mobilized  (n=171) | Mobilized  (n=328) | P value |
| Total comorbidities (IQR) | 2 (3) | 2 (2) | 0.089 |
| Currently receiving chemotherapy  Congenital anomalies  Congestive Heart Failure  Smoker  Renal failure  Prior cerebral vascular accident  Diabetes  Disseminated cancer  Advanced directive limiting care  History of angina  Prior myocardial infarction  Peripheral vascular disease  Hypertension  Prematurity  COPD  Steroid use  Cirrhosis  Other major non-psychiatric disorder  EtOH use disorder  Other substance use disorder  Dementia  ADHD  Other major psychiatric disorder | 3 (1.75%)  1 (0.58%)  18 (10.53%)  53 (30.99%)  15 (8.77%)  17 (9.94%)  32 (18.71%)  12 (7.02%)  42 (24.56%)  0 (0%)  10 (5.85%)  11 (6.43%)  61 (35.67%)  0 (0%)  11 (6.43%)  0 (0%)  8 (4.68%)  2 (1.17%)  25 (14.62%)  29 (16.96%)  7 (4.09%)  3 (1.75%)  33 (19.30%) | 2 (0.61%)  1 (0.30%)  34 (10.37%)  83 (25.30%)  30 (9.15%)  31 (9.45%)  51 (15.55%)  18 (5.49%)  21 (6.40%)  1 (0.30%)  21 (6.40%)  17 (5.18%)  143 (43.60%)  0 (0%)  24 (7.32%)  5 (1.52%)  4 (1.22%)  2 (0.61%)  34 (10.37%)  39 (11.89%)  11 (3.35%)  6 (1.83%)  79 (24.09%) | 0.22  0.64  0.96  0.18  0.89  0.86  0.37  0.50  <0.0001  0.47  0.81  0.56  0.087  -  0.71  0.10  0.017  0.51  0.16  0.12  0.67  0.95  0.22 |

| **Supplemental Table 4. Complications after-COVID stratified by mobilization** | | | |
| --- | --- | --- | --- |
| Complications (%) | Not Mobilized  (n= 171) | Mobilized  (n= 328) | P value |
| Total complications (IQR) | 0 (1) | 0 (1) | 0.5775 |
| Deep surgical infection  Drug/EtOH substance use disorder  Deep venous thrombosis (DVT)  Compartment syndrome  Graft/ prosthesis/ flap failure  Myocardial infarction  Organ space infection  Osteomyelitis  Pneumonia  Pulmonary embolism  Sepsis  Stroke/cerebral vascular accident  Superficial infection  Urinary tract infection | 0 (0%)  2 (1.17%)  8 (4.68%)  0 (0%)  0 (0%)  2 (1.17%)  0 (0%)  0 (0%)  20 (11.70%)  0 (0%)  11 (6.43%)  6 (3.51%)  0 (0%)  2 (1.17%) | 5 (1.52%)  6 (1.83%)  13 (3.96%)  2 (0.61%)  1 (0.30%)  0 (0%)  2 (0.61%)  1 (0.30%)  38 (11.59%)  7 (2.13%)  23 (7.01%)  6 (1.83%)  5 (1.52%)  12 (3.66%) | 0.10  0.58  0.71  0.31  0.47  0.05  0.31  0.47  0.97  0.054  0.81  0.25  0.10  0.11 |
